# A clinicoradiological model for preoperative prediction of lateral lymph node metastasis in rectal cancer

**DOI:** 10.64898/2026.04.13.26350816

**Authors:** Qiyuan Shen, Guancong Wang, Muhai Fu, Kaiyuan Yao, Qunzhang Zeng, Yugang Yang, Yincong Guo

## Abstract

**Background:** Lateral lymph node metastasis (LLNM) is associated with poor prognosis in patients with rectal cancer and may influence the indication for lateral lymph node dissection. Accurate preoperative identification of LLNM remains challenging. This study aimed to develop and internally validate a clinicoradiological model for preoperative prediction of LLNM in rectal cancer.

**Methods:** A retrospective cohort of 64 patients undergoing lateral lymph node dissection (LLND) for rectal cancer was analysed; 21 (32.8%) had pathological lateral lymph node metastasis (LLNM). A prespecified preoperative clinicoradiological model was fitted using penalised logistic regression with L2 regularisation (ridge), incorporating MRI-measured lateral lymph node short-axis diameter (LLN-SAD), dichotomised clinical T stage (T3-4 vs T1-2), dichotomised clinical N stage (N+ vs N0), and log(CA19-9+1). Model performance was evaluated using the area under the receiver operating characteristic curve (AUC), calibration analysis, and bootstrap internal validation.

**Results:** The model showed good discrimination (AUC 0.914), with an optimism-corrected AUC of 0.887 on bootstrap validation. Calibration remained acceptable after optimism correction (calibration intercept −0.127; slope 1.045). Decision curve analysis suggested net benefit across clinically relevant threshold probabilities, particularly between 0.10 and 0.30. The model was implemented as a web-based calculator to facilitate clinical use.

**Conclusion:** This clinicoradiological model showed good discrimination, acceptable calibration, and potential clinical utility for preoperative assessment of LLNM risk in rectal cancer. It may assist individualized risk stratification and treatment planning, although external validation is required before routine clinical implementation.

## Background

The management of rectal cancer took on new clinical urgency in our practice following a case of severe postoperative complications after lateral lymph node dissection (LLND), prompting the development of a robust predictive model to rigorously evaluate surgical selection criteria—particularly given the generally low positivity rates of this procedure.

Colorectal cancer ranks among the most prevalent malignancies globally, with therapeutic strategies heavily contingent upon tumor staging and nodal involvement. Lateral lymph node metastasis (LLNM) has emerged as a pivotal prognostic determinant in locally advanced rectal cancer, attracting intense scrutiny due to its distinctive anatomical challenges and ongoing therapeutic controversies [1].Current guidelines reflect geographical disparities: Western protocols (e.g., NCCN) prioritize neoadjuvant chemoradiation followed by total mesorectal excision (TME), whereas East Asian approaches (notably Japan) advocate prophylactic LLND for high-risk cases. The divergence stems from differing perceptions regarding the risks of lymph node metastasis and radiosensitivity [2].

In China, LLNM is documented in 10-20% of low rectal cancers, correlating significantly with increased locoregional recurrence and diminished survival [3]. Reliable preoperative identification of high-risk patients remains elusive. While MRI-based assessment of nodal morphology (short-axis diameter, border irregularity) constitutes the diagnostic mainstay, but assessing border irregularity is difficult to quantify and heavily dependent on radiologists’ experience, resulting in substantial subjectivity [4].An imaging-driven multivariable predictive model could provide a more precise reference for clinical decision-making.

This study synthesized clinical and radiological characteristics to identify independent predictive factors for LLNM, providing an evidence-based foundation for optimizing therapeutic decision-making in rectal cancer patients(Figure 1).

**Figure 1.**
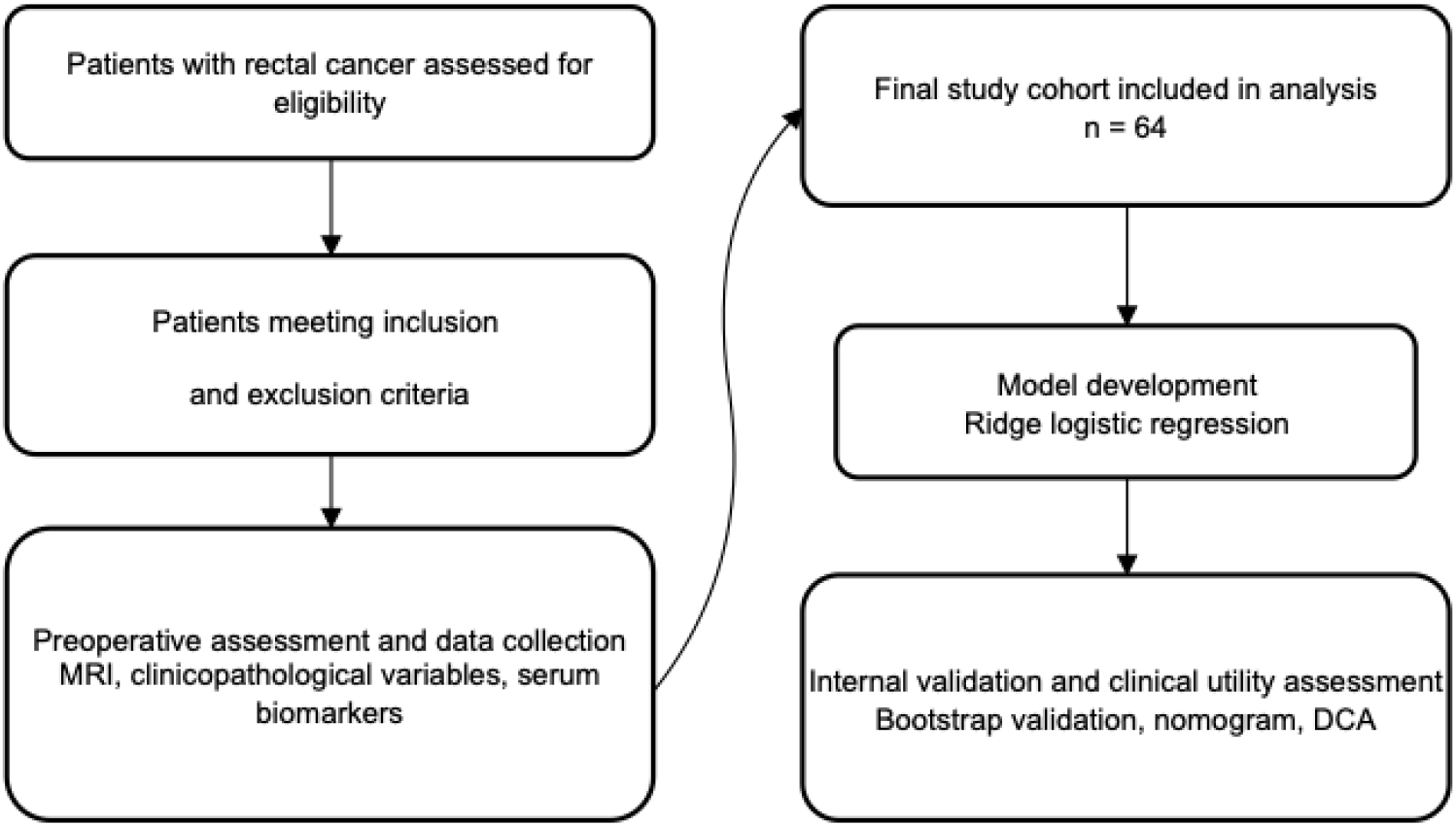
Study flowchart

## Materials and Methods

### Study design and patients

This retrospective cohort study was reported in accordance with the STROBE guidelines. Consecutive patients with rectal cancer who underwent radical surgery consisting of total mesorectal excision (TME) with lateral lymph node dissection (LLND) at the Department of Colorectal and Anal Surgery, Zhangzhou Affiliated Hospital of Fujian Medical University, between January 2019 and June 2023 were screened for eligibility. Patients were eligible if they had pathologically confirmed primary rectal adenocarcinoma, a tumour located within 10 cm from the anal verge on MRI, a lateral lymph node short-axis diameter of at least 4 mm on pretreatment MRI, and had completed baseline rectal MRI before initiation of any treatment, including surgery or neoadjuvant therapy. Exclusion criteria were a history of prior pelvic surgery, concurrent malignancies, synchronous unresectable distant metastases, or unavailable pretreatment MRI data. Consecutive patients were included to minimize selection bias and improve the representativeness of the study population. Written informed consent was obtained from all participants, and the study protocol was approved by the institutional ethics committee.

### Surgical Procedures

All patients underwent standard TME combined with LLND adhering to the anatomical principle of “two planes and three tracts”: The peritoneal dissection continued along the external iliac artery and common iliac artery, followed by opening the vascular sheaths of the external iliac artery and vein. The surrounding lymphatic fatty tissue was en bloc mobilized medialward until the bifurcation of the internal and external iliac arteries was identified. Systematic skeletonization was then performed along the internal iliac artery trunk and its branches (superior gluteal artery, obturator artery, etc.), with particular attention to preserving the internal iliac vein and its tributaries. The obturator fascia was opened superficial to the obturator nerve, and dissection proceeded along the nerve course while carefully identifying and protecting the obturator neurovascular bundle. The surgical field after completion of right-sided LLND is demonstrated (Figure 2).

**Figure.2.**
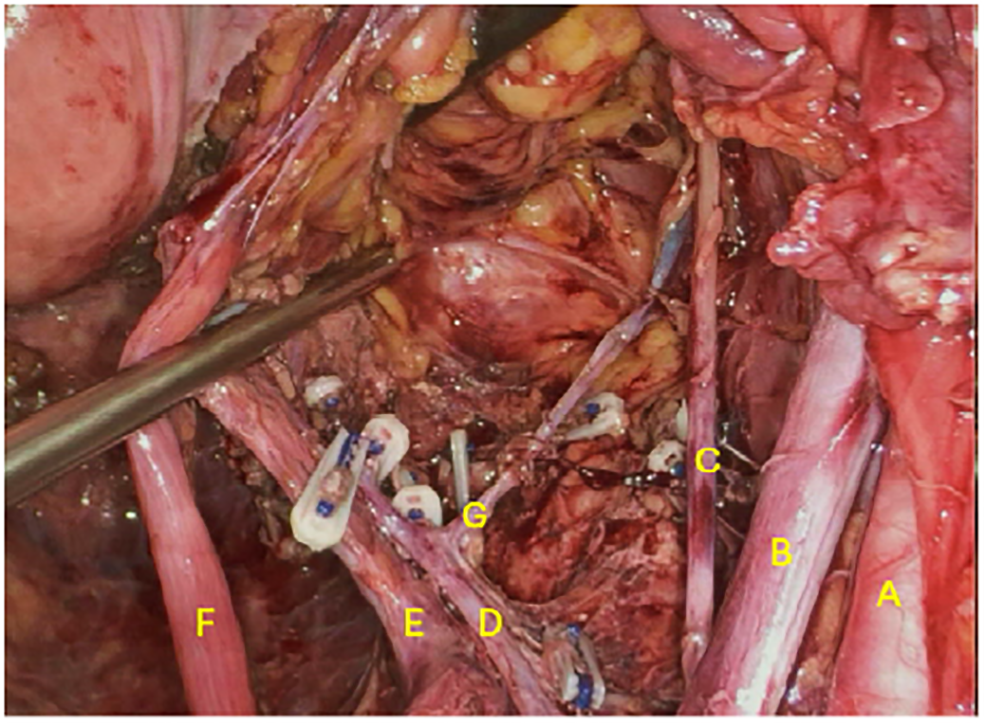
Illustrates the surgical field after right lateral lymph node dissection. Labeled structures include: A, external iliac artery; B, external iliac vein; C, obtutator nerve; D, internal iliac artery, E, internal iliac vein; F, ureter; G, obturator artery.

### Radiological Evaluation and Grouping Criteria

The depth of tumor invasion (MRI-based cT) and mesorectal lymph node status (MRI-based cN) were evaluated by radiologists based on preoperative MRI findings. Patients were subsequently stratified into two groups based on postoperative pathological findings: lateral lymph node metastasis-positive (LLNM+) or negative (LLNM-).

### Statistical Analysis

Continuous variables were summarized as mean ± standard deviation or median with interquartile range, depending on distribution normality assessed by the Shapiro–Wilk test. Categorical variables were presented as counts. Comparisons between patients with and without lateral lymph node metastasis (LLNM) were performed using the independent-samples t test or Mann–Whitney U test for continuous variables, and Fisher’s exact test for categorical variables. All statistical analyses were performed using SPSS 26.0 (IBM) and R software (version 2.15.3). A two-sided P value <0.05 was considered statistically significant.

### Use of Artificial Intelligence (AI) Tools

During the preparation of this manuscript, the authors used ChatGPT (OpenAI, San Francisco, CA, USA) solely for language translation and linguistic editing to improve clarity and readability. The AI tool was not used for data analysis, study design, interpretation of results, or generation of scientific content. All scientific content, conclusions, and interpretations were independently reviewed and verified by the authors, who take full responsibility for the integrity and accuracy of the manuscript.

### Model development

#### Candidate predictors

Candidate predictors were prespecified based on clinical relevance, preoperative availability, and prior evidence, rather than univariate significance testing. Given the limited number of LLNM events, model complexity was restricted to minimize overfitting.

#### Predictor specification

The primary model included LLN short-axis diameter on MRI (LLN-SAD), clinical T stage, clinical N stage, and serum CA19-9. T stage was dichotomised (T1–2 vs T3–4) and N stage was dichotomized (N0 vs N+ ) to reduce degrees of freedom. Continuous variables were retained on their original scale where possible; CA19-9 was log-transformed to address right skew (log[CA19-9 + 1]).

#### Additional variables

Age, sex, tumour distance from the anal verge, tumour longest diameter, differentiation grade, serum CEA, and laterality were summarised descriptively but not included in the primary model owing to limited incremental value and the need for parsimony. These factors were considered for exploratory or sensitivity analyses. These decisions were made a priori to balance predictive performance, interpretability, and model stability.

#### Nomogram derivation

For nomogram presentation, an unpenalised logistic model was uniformly shrunken using the bootstrap-derived calibration slope, with intercept recalibration using an offset approach.

#### Model performance and validation

Penalised logistic regression with L2 regularisation (ridge) was fitted, with the penalty parameter selected by 10-fold cross-validation. Discrimination was assessed using the area under the receiver operating characteristic curve (AUC). Internal validation used bootstrap resampling to obtain optimism-corrected calibration (calibration plots, intercept and slope) and Brier score. Clinical utility was evaluated using decision curve analysis across clinically relevant threshold probabilities. For nomogram presentation, coefficients from an unpenalised logistic model were uniformly shrunken using the bootstrap-derived calibration slope, with intercept recalibration using an offset approach.

## Results

### Study population

The study flow is shown in Figure 1. To ensure cohort homogeneity, 10 patients who underwent LLND for recurrent disease after previous radical resection were excluded (including 5 rectal cancers, 1 sigmoid cancer and 4 with missing data). A total of 64 patients undergoing LLND were included; 21 (32.8%) had pathological LLNM.

### Baseline Characteristics

Baseline characteristics are summarised in Table 1. Compared with the LLNM− group (n=43), the LLNM+ group (n=21) had a larger MRI-measured LLN short-axis diameter (LLN-SAD; median 10.0 (i.q.r. 9.0–14.0) vs 8.0 (6.5–9.0) mm; P<0.001), and more frequently had clinical T3–4 disease (90.5% vs 53.5%; P=0.003) and clinical N+ disease (85.7% vs 32.6%; P<0.001), with higher CA19-9 levels (median 21.5 (17.6–72.0) vs 12.0 (6.5–25.1); P=0.004).

**Table 1.** Baseline Characteristics Stratified by Lateral Lymph Node Metastasis Status.

| Variable | Overall (n=64) | LLNM+ (n=21) | LLNM– (n=43) | P-value |
| --- | --- | --- | --- | --- |
| Age (years) | 62.8 ± 10.8 | 64.4 ± 12.6 | 62.1 ± 9.9 | **0.467** |
| Gender |  |  |  | **0.127** |
| Male | 39 (60.9) | 10 (47.6) | 29 (67.4) |  |
| Female | 25(39.1) | 11 (52.4) | 14 (32.6) |  |
| Tumor-anal verge<sup>a</sup> (mm) | 57.5 [40.0–70.0] | 70.0 [30.0–76.0] | 54.0 [40.5–61.5] | **0.360** |
| LLN-SAD<sup>b</sup> (mm) | 8.0 [7.0–10.0] | 10.0 [9.0–14.0] | 8.0 [6.5–9.0] | **0.001** |
| CEA | 5.0 [2.7–11.9] | 5.0 [2.9–7.1] | 5.4 [2.7–13.8] | **0.858** |
| CA19-9 | 15.1 [8.5–39.0] | 21.5 [17.6–72.0] | 12.0 [6.5–25.1] | **0.004** |
| T stage |  |  |  | **0.003** |
| 1-2 | 22 (34.4) | 2 (9.5) | 20 (46.5) |  |
| 3-4 | 42 (65.6%) | 19 (90.5) | 23 (53.5) |  |
| N stage |  |  |  | **<0.001** |
| 0 | 32 (50.0) | 3 (14.3); | 29 (67.4) |  |
| + | 32 (50.0) | 18 (85.7) | 14 (32.6) |  |
| Tumor Differentiation |  |  |  | **0.507** |
| - Well / Moderate | 52 (81.2) | 16 (76.2) | 36 (83.7) |  |
| - Poor | 12 (18.8) | 5 (23.8) | 7 (16.3) |  |
| **Tumor Longest Diameter** | 54.0 [43.0–70.2] | 56.0 [43.0–75.0] | 52.0 [42.5–68.5] | **0.457** |
| **Location of dissected lymph nodes** |  |  |  | **0.254** |
| - Left | 38 ( 59.4 ) | 12 (57.1) | 26 (60.5) |  |
| - Right | 26 ( 40.7 ) | 9 (42.9) | 17(39.5) |  |

**Note:**
**a Tumor-anal verge distance indicates the measurement from the tumor to the anal verge.**
**b LLN-SAD, lateral lymph node short-axis diameter measured on MRI.**

### Regression analyses

Univariable and multivariable logistic regression analyses are presented in Table 2. LLN-SAD (OR 1.44 per mm, 95% CI 1.16 to 1.79; P=0.001), clinical T stage (OR 8.26, 1.71 to 39.92; P=0.009), clinical N stage (OR 12.43, 3.13 to 49.34; P<0.001) and log(CA19-9+1) (OR 2.12, 1.21 to 3.72; P=0.009) were associated with LLNM on univariable analysis. In the prespecified multivariable model, LLN-SAD (OR 1.50 per mm, 1.12 to 2.01; P=0.007), clinical T stage (OR 9.64, 1.18 to 78.42; P=0.034) and clinical N stage (OR 11.52, 2.11 to 62.89; P=0.005) remained independently associated, whereas the association for log(CA19-9+1) attenuated (OR 1.87, 0.93 to 3.73; P=0.077).

**Table 2.** Univariable and multivariable logistic regression analyses of predictors associated with LLNM.

| Variable | Univariate |  | Multivariable |  |
| --- | --- | --- | --- | --- |
|  | OR (95% CI) | P | OR (95% CI) | P |
| **LLN-SAD** | 1.44 (1.16–1.79) | 0.001 | 1.50 (1.12–2.01) | 0.007 |
| **T stage** | 8.26 (1.71–39.92) | 0.009 | 9.64 (1.18–78.42) | 0.034 |
| **N stage** | 12.43 (3.13–49.34) | <0.001 | 11.52 (2.11–62.89) | 0.005 |
| **log(CA19-9 + 1)** | 2.12 (1.21–3.72) | 0.009 | 1.87 (0.93–3.73) | 0.077 |
| **CEA** | 1.01 (1.00–1.02) | 0.243 | — | — |
| **Age** | 1.02 (0.97–1.07) | 0.420 | — | — |
| **Male sex** | 0.44 (0.15–1.28) | 0.131 | — | — |
| **Tumour distance from anal verge** | 1.00 (0.98–1.02) | 0.991 | — | — |
| **Tumour longest diameter** | 1.01 (0.99–1.03) | 0.381 | — | — |
| **Differentiation** | 0.62 (0.17–2.26) | 0.471 | — | — |
| **Side** | 1.15 (0.40–3.31) | 0.799 | — | — |

### Model presentation

The final model was presented as a nomogram and implemented as an online calculator for individualized preoperative risk estimation (Figure 3). The predictive model was constructed using multivariable logistic regression as follows: logit(P) = -9.5683 + 0.4044 × [LLN-SAD] + 2.2656 × [T stage] + 2.4440 × [N stage] + 0.6237 × log(CA19-9 + 1),where P denotes the probability of lateral lymph node metastasis, the predicted probability was calculated as: P = 1 / (1 + exp(-logit(P))).

**Figure 3.**
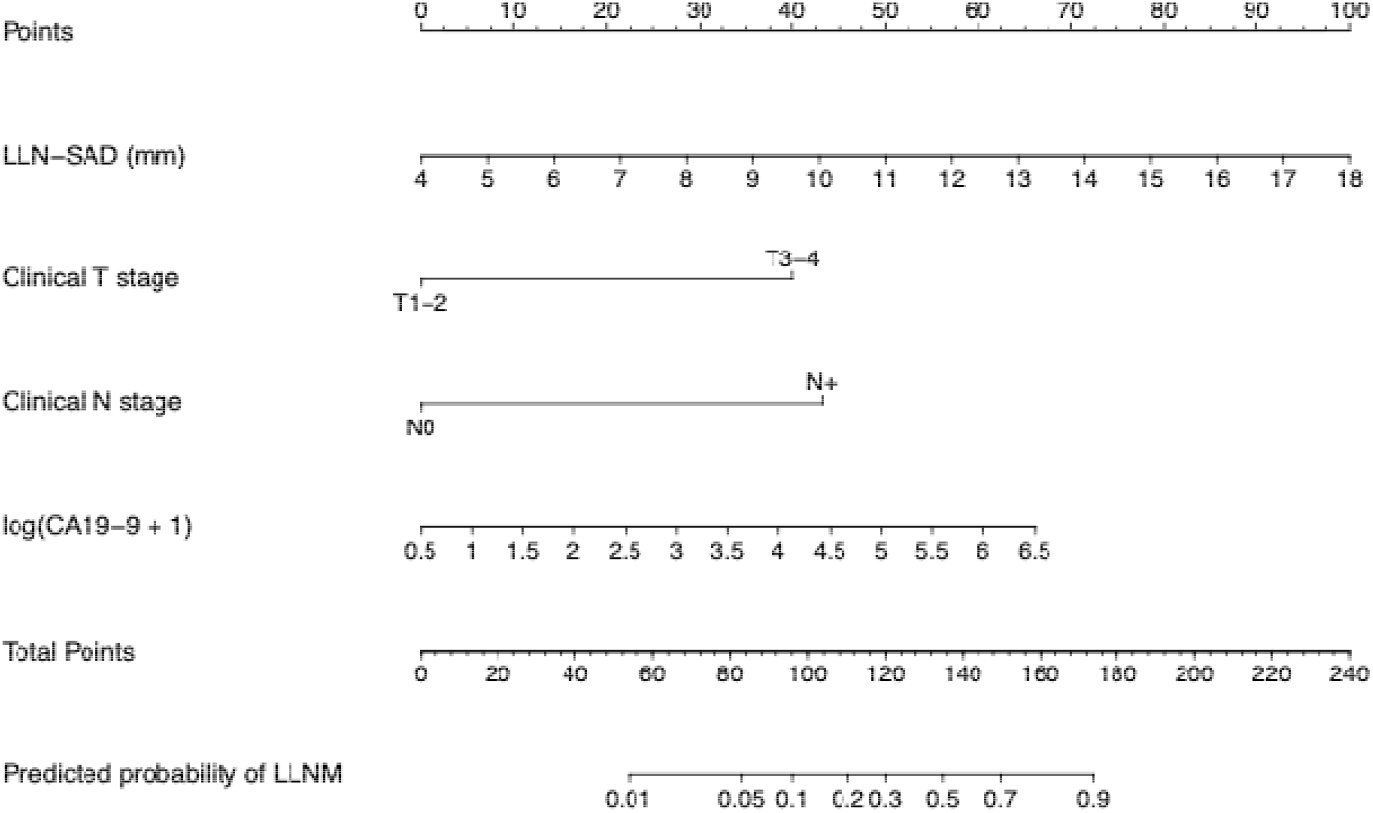
Nomogram. Nomogram for predicting lateral lymph node metastasis in patients with rectal cancer, incorporating LLN-short axis diameter on MRI (LLN SAD), clinical T stage, clinical N stage, and serum CA 19-9 [log(CA19-9 +1)]. For categorical variables, categories located towards the right side of the scale correspond to higher predicted risk.

An online calculator based on the present model is publicly available at:

https://rectal-llnm-risk.shinyapps.io/Rectal-LLNM-Risk-Calculator/

### Diagnostic performance

At the Youden-optimal threshold, the multivariable model showed the best overall diagnostic performance (Youden index 0.742; sensitivity 0.905; specificity 0.837; accuracy 0.859; NPV 0.947) compared with individual predictors (Table 3). Thresholds were derived for descriptive comparison and were not used for variable selection; clinical decision thresholds were evaluated using decision curve analysis.

**Table 3.** Optimal cut-offs (Youden index) and diagnostic performance of individual predictors and the multivariable model for LLNM.

| Predictor | Cut-off (Youden) | Youden index | Sensitivity | Specificity | Accuracy | PPV | NPV |
| --- | --- | --- | --- | --- | --- | --- | --- |
| LLN-SAD | $\geq 8.75$ mm | 0.460 | 0.762 | 0.698 | 0.719 | 0.552 | 0.857 |
| T stage (T3–4 vs T1–2) | 1 vs 0 | 0.370 | 0.905 | 0.465 | 0.609 | 0.452 | 0.909 |
| N stage (N+ vs N0) | 1 vs 0 | 0.532 | 0.857 | 0.674 | 0.734 | 0.562 | 0.906 |
| log(CA19-9+1) | $\geq 2.86$ | 0.436 | 0.762 | 0.674 | 0.703 | 0.533 | 0.853 |
| Multivariable model | $\geq 0.998$ | 0.742 | 0.905 | 0.837 | 0.859 | 0.731 | 0.947 |

### Model performance, calibration and clinical utility

The model showed good discrimination (AUC 0.914; Figure 4). Bootstrap internal validation suggested limited optimism, with an optimism-corrected AUC of 0.887; calibration remained acceptable (calibration intercept −0.127; slope 1.045). The optimism-corrected calibration curve closely followed the ideal reference line, indicating adequate calibration after internal validation. (Figure 6).

**Figure 4.**
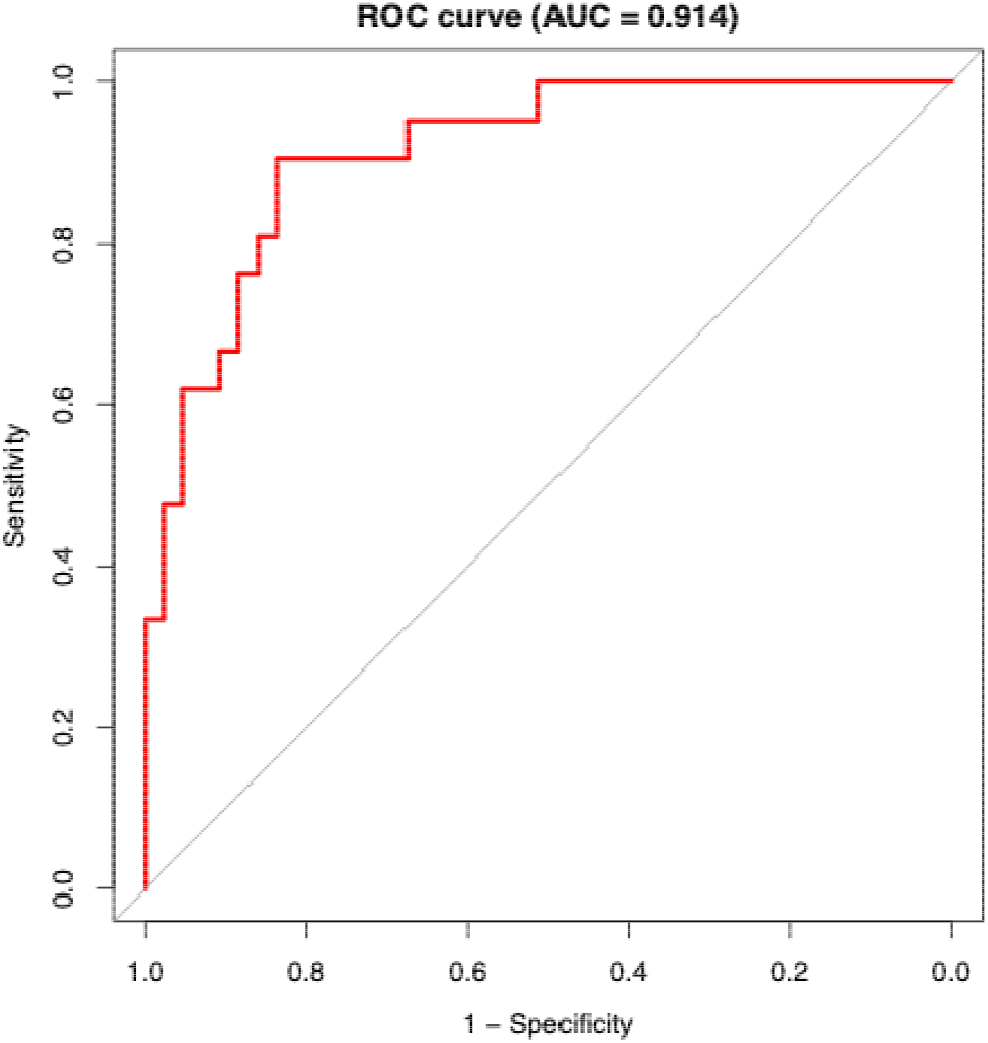
ROC curve. Receiver operating characteristic (ROC) curve of the nomogram model for predicting lateral lymph node metastasis.

### Decision curve analysis

Decision curve analysis demonstrated that the nomogram provided greater net benefit than the “treat-all” and “treat-none” strategies across a range of clinically relevant threshold probabilities, particularly between 0.10 and 0.30 (Figure 7). The multivariable model outperformed each individual predictor on AUC comparison (Figure 5; Table 4).

**Table 4.** Comparison of AUCs between the multivariable model and individual predictors using DeLong’s test.

| Comparison | AUC_Multi | AUC_Uni | Z_Score | p_Value |
| --- | --- | --- | --- | --- |
| Multivariate vs LLN-SAD | 0.914 | 0.762 | 1.056 | 0.291 |
| Multivariate vs T stage | 0.914 | 0.685 | 1.945 | 0.052 |
| Multivariate vs N stage | 0.914 | 0.766 | 1.370 | 0.171 |
| **Multivariate vs log(CA19-9+1)** | 0.914 | 0.721 | 1.177 | 0.239 |

**Figure.5.**
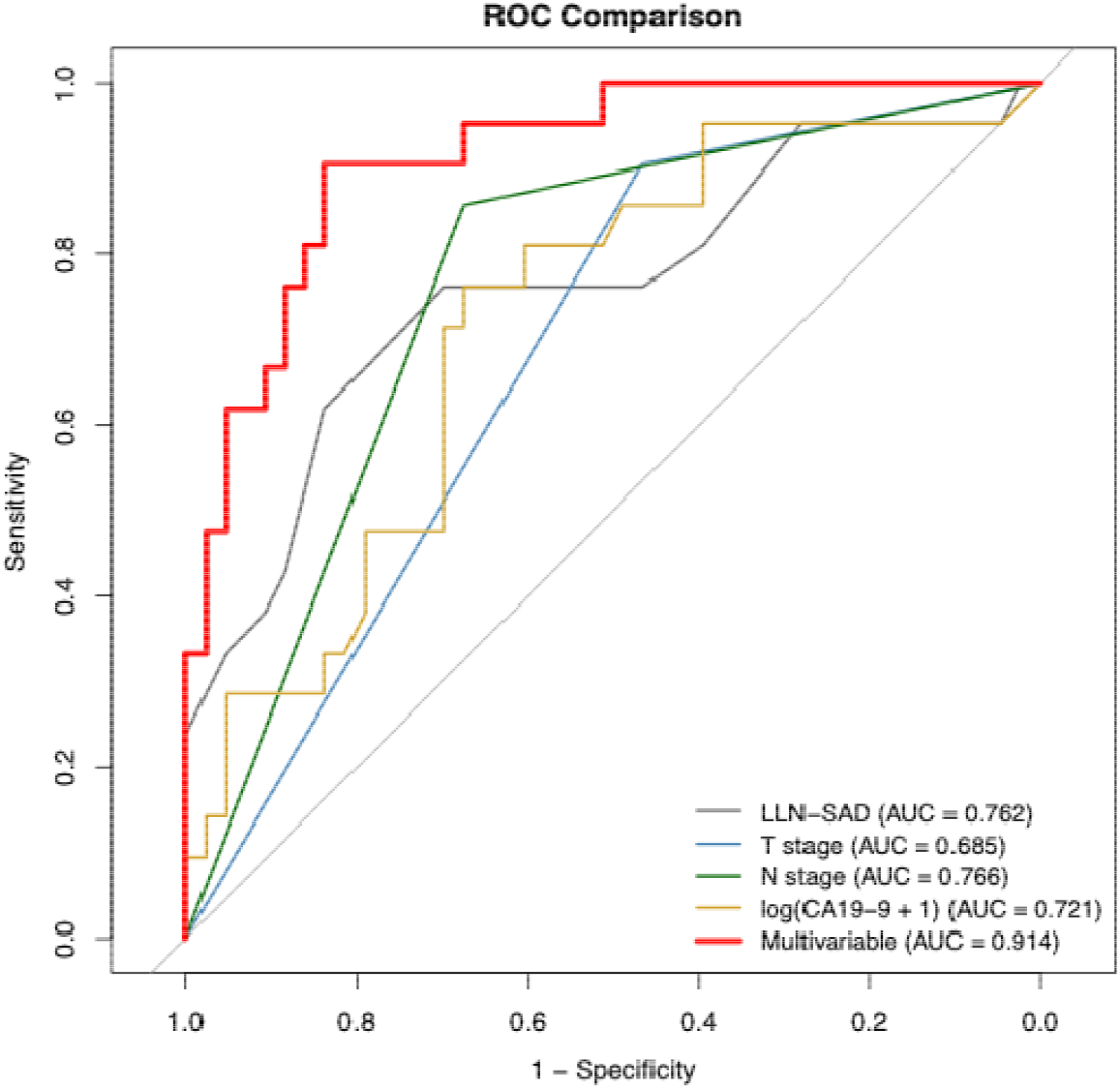
ROC comparison. Receiver operating characteristic curves comparing the discriminative performance of individual predictors and the multivariable nomogram model.

**Figure 6.**
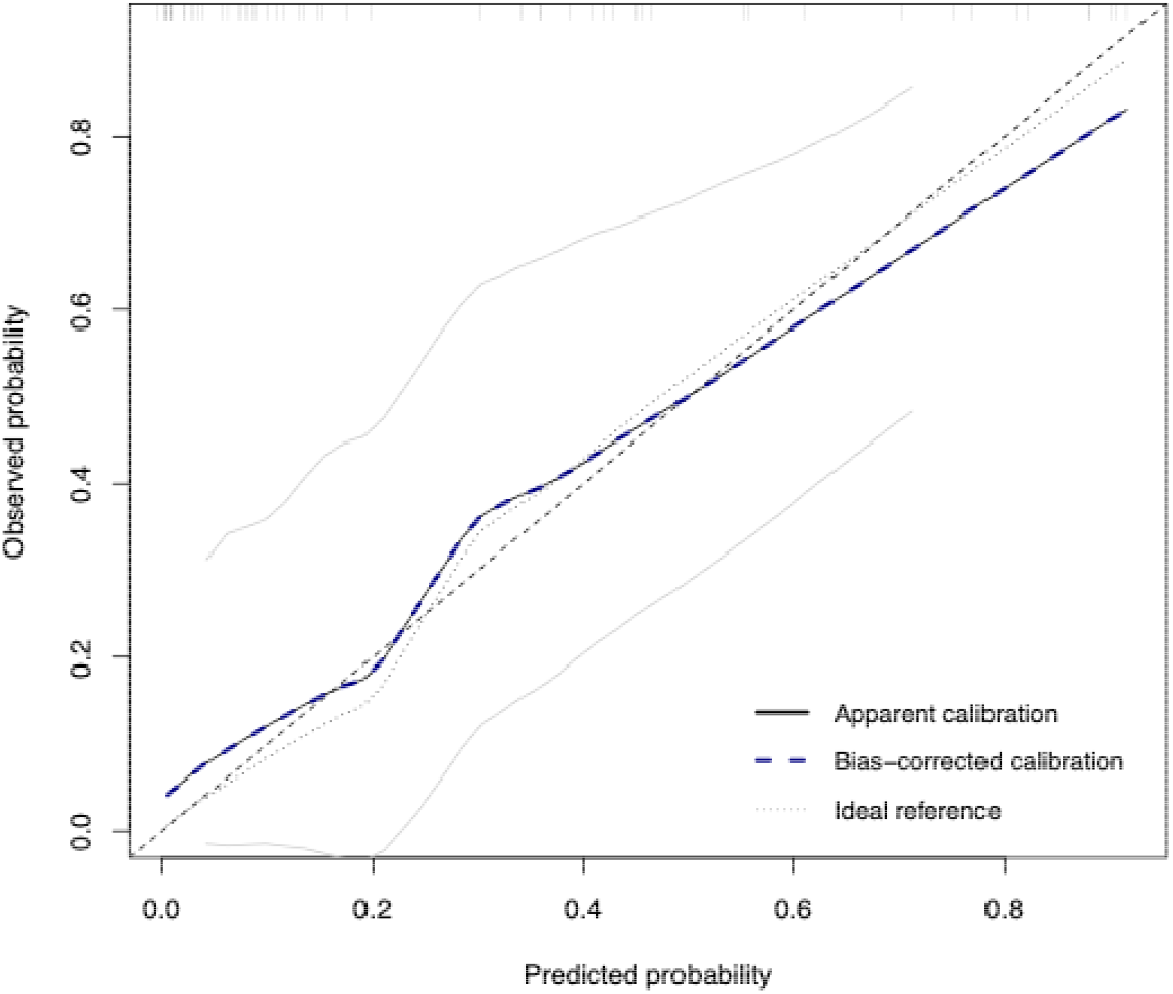
Calibration plot. Calibration plot of the nomogram model. The dashed line represents perfect calibration, the solid line shows the LOESS-smoothed relationship between predicted and observed risks, and points Indicate observed event rates within deciles of predicted probability.

**Figure 7.**
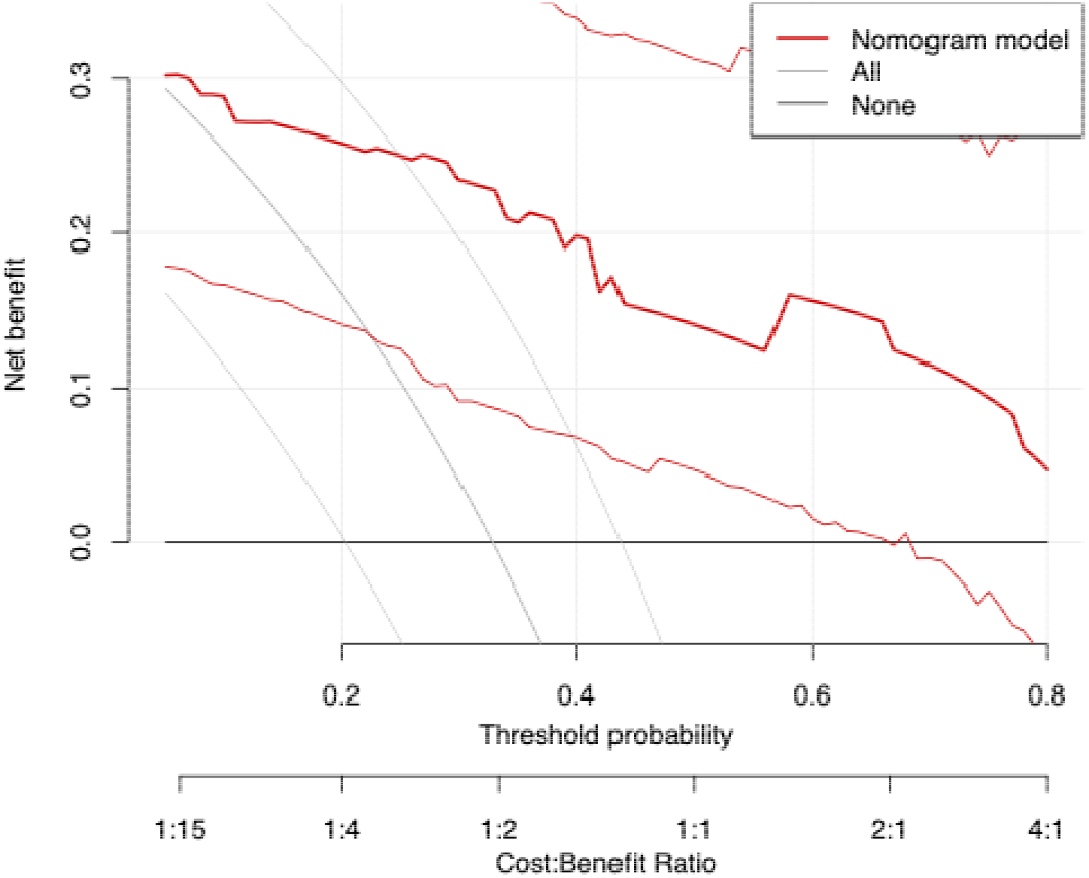
Decision curve analysis. Decision curve analysis demonstrating the net benefit of the nomogram compared with the “treat-all” and “treat-none’ strategies across a range of threshold probabilities.

## Discussion

### Principal findings

This study developed and internally validated a clinicoradiological prediction model for pathological lateral lymph node metastasis (LLNM) in patients undergoing lateral lymph node dissection. Using four prespecified preoperative predictors (LLN-SAD, clinical T stage, clinical N stage and log(CA19-9+1)), the model showed good discrimination and acceptable bootstrap-corrected calibration, with clinically relevant net benefit on decision curve analysis.

### Interpretation of predictors

LLN short-axis diameter on MRI (LLN-SAD) was a consistent predictor, supporting the importance of nodal morphology and size in preoperative risk stratification [1,3,4]. Unlike fixed size thresholds, LLN-SAD was retained as a continuous measure in the primary model to preserve prognostic information and avoid loss of discrimination. Clinical T3–4 stage and mesorectal nodal positivity (N+) also remained independently associated with LLNM [4], consistent with more advanced local disease being linked to a higher likelihood of extramesorectal spread.

The association of CA19-9 attenuated after adjustment for imaging and stage variables, suggesting partial overlap in predictive information rather than absence of clinical relevance.

### Clinical implications

Previous studies have suggested that risk-stratified approaches are critical in managing LLNM. For patients assessed preoperatively as having high-risk LLNM, more aggressive therapeutic strategies may be considered, including neoadjuvant chemoradiotherapy or standardized LLND [5]; For low-risk patients, conservative management without prophylactic lymph node dissection may be preferable to avoid treatment-related morbidity [6].

Our study demonstrates that the integration of radiological and pathological biomarkers achieves satisfactory predictive performance for LLNM risk stratification. The model provides an interpretable tool for individualised preoperative risk estimation. Decision curve analysis indicated that use of the model may be most informative within threshold probabilities of approximately 0.10–0.30, a range in which decisions regarding intensified neoadjuvant therapy, selective LLND, or closer surveillance are often most uncertain [7-10]. The nomogram presentation facilitates bedside use, while the underlying model performance should guide interpretation.

### Methodological considerations

Given the limited number of LLNM events, model complexity was deliberately restricted and penalised logistic regression with L2 regularisation (ridge) was used to stabilise estimates. Internal validation used bootstrap resampling to quantify optimism and to support coefficient shrinkage with intercept recalibration for the nomogram presentation. After internal validation, the multivariable model retained strong discriminative ability with minimal optimism, suggesting that its performance gain was not merely attributable to overfitting. In contrast, while certain single predictors such as N-stage or LLN-SAD demonstrated reasonable standalone discrimination, their performance was less stable and provided limited clinical decision support when used in isolation.

### Tumour level and nodal size thresholds

Tumour distance from the anal verge was not associated with LLNM in the present cohort (P=0.991). This contrasts with anatomical and clinical observations that low rectal tumours (≤5 cm from the anal verge) may preferentially spread via lateral lymphatic pathways to the internal iliac and obturator basins, informing contemporary indications for LLND in low rectal cancer and in the presence of radiologically suspicious lateral nodes on MRI (short-axis ≥7 mm) [7-10]. A plausible explanation for the null association observed here is restricted variability in tumour height, as the study population comprised patients selected for LLND on the basis of preoperative assessment and suspected nodal disease, which may enrich for low tumours and narrow the distribution of anal verge distance [11,12]. This highlights the need for external validation in less selected cohorts and supports further evaluation of tumour-level–specific nodal size thresholds.

### Limitations

Several limitations merit consideration. The retrospective single-centre design and inclusion of patients undergoing LLND may limit generalisability to broader rectal cancer populations. Imaging measurements may vary across scanners and readers, and external validation in independent cohorts is required before routine implementation.

### Imaging and biological heterogeneity

Advanced imaging may further refine preoperative risk stratification. PET-CT, where available, could be evaluated as an adjunct to pelvic MRI to improve detection of metabolically active nodal disease, particularly in equivocal cases [8]. In addition, the biology of metachronous or recurrent lateral nodal disease may differ from that of primary LLNM. Future work should explore whether histopathological features (for example extramural venous invasion or poor differentiation), surgical technique, and response to neoadjuvant therapy contribute to late nodal relapse, and whether separate risk models are warranted for primary versus recurrent cohorts.

### Future work

Future work should prioritise prospective multicentre validation, assessment of interobserver reproducibility, and evaluation of whether advanced imaging sequences and radiomic or deep-learning features can improve detection of micrometastatic disease and enhance generalisability across clinical settings.

## Conclusion

This clinicoradiological model showed good discrimination and acceptable calibration for preoperative prediction of lateral lymph node metastasis in rectal cancer. The model, together with its web-based calculator, may assist individualized preoperative risk stratification and treatment planning. However, external validation is required before routine clinical use.

## Data Availability

All data produced in the present study are available upon reasonable request to the authors
All data produced in the present work are contained in the manuscript

## Author contributions

Protocol/project development: Qiyuan Shen, Yincong Guo; Data collection or management: Muhai Fu, Yugang Yang, Qunzhang Zeng; Data analysis: Qiyuan Shen, Guancong Wang; Manuscript writing/editing and prepared figures: Qiyuan Shen. All authors reviewed the manuscript.

## Data availability

All data obtained or analyzed during this work are included within the article.

